## Supplementary figures and images for "Evidence for Virus-Mediated Oncogenesis in Bladder Cancers Arising in Solid Organ Transplant Recipients"

### Supplemental Figures

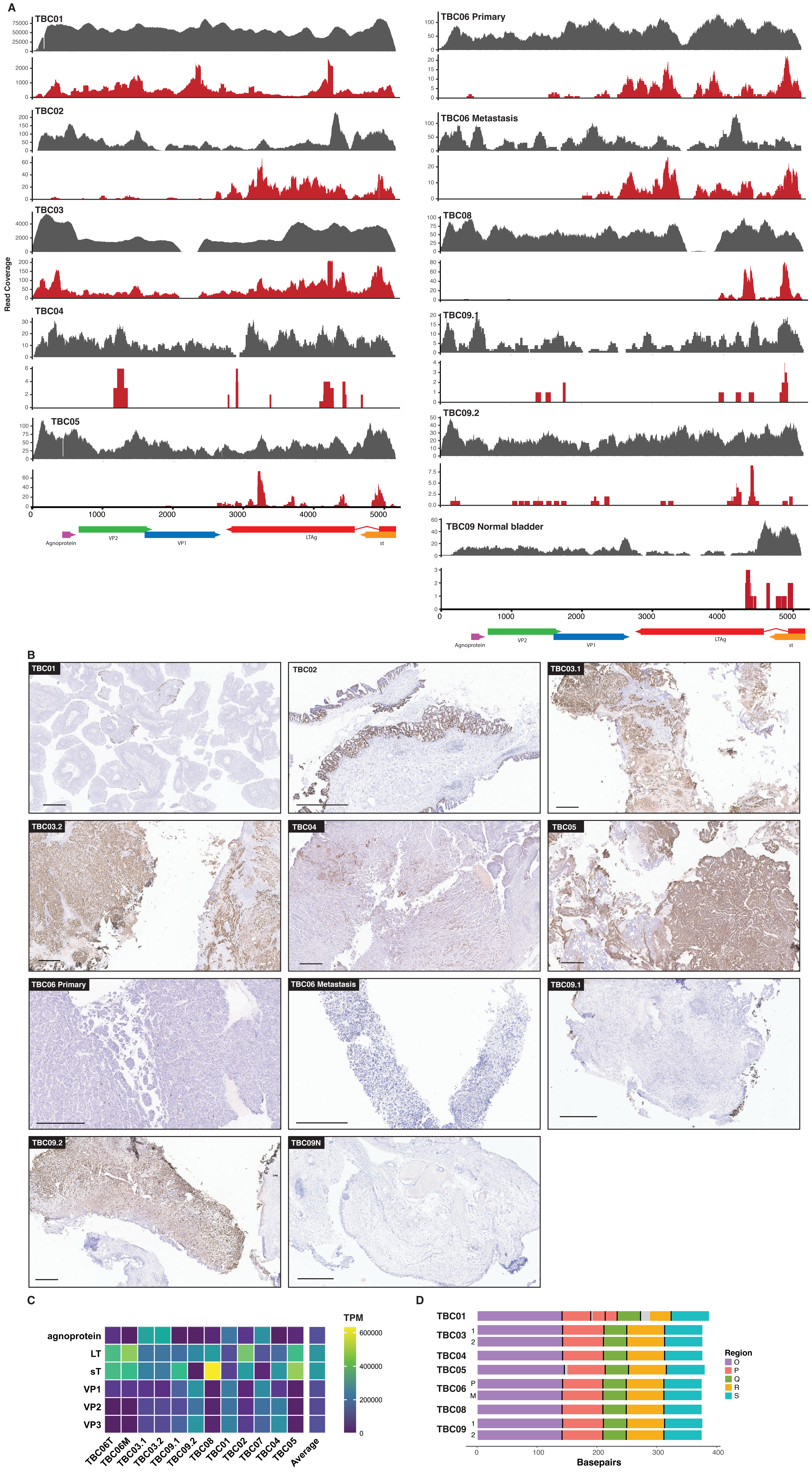

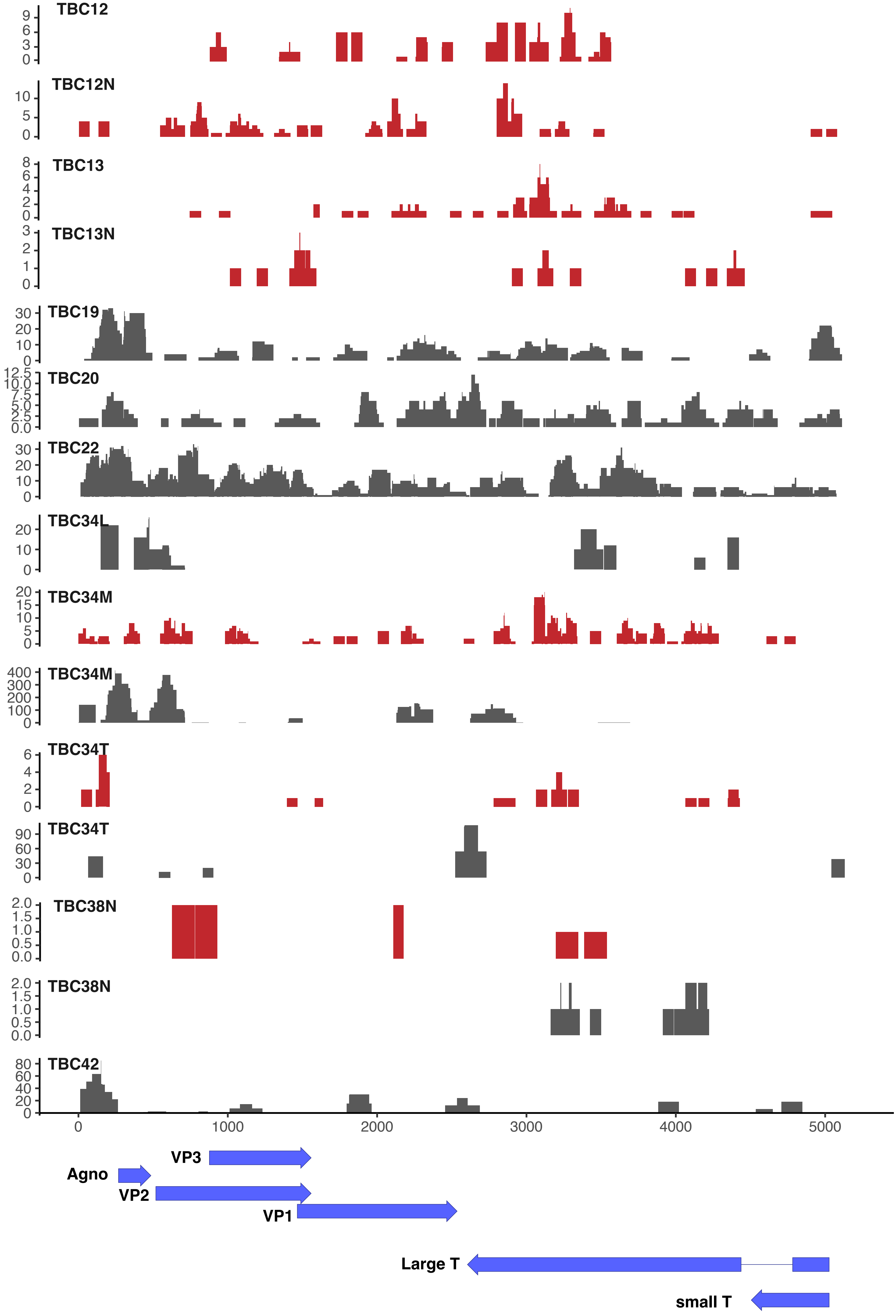

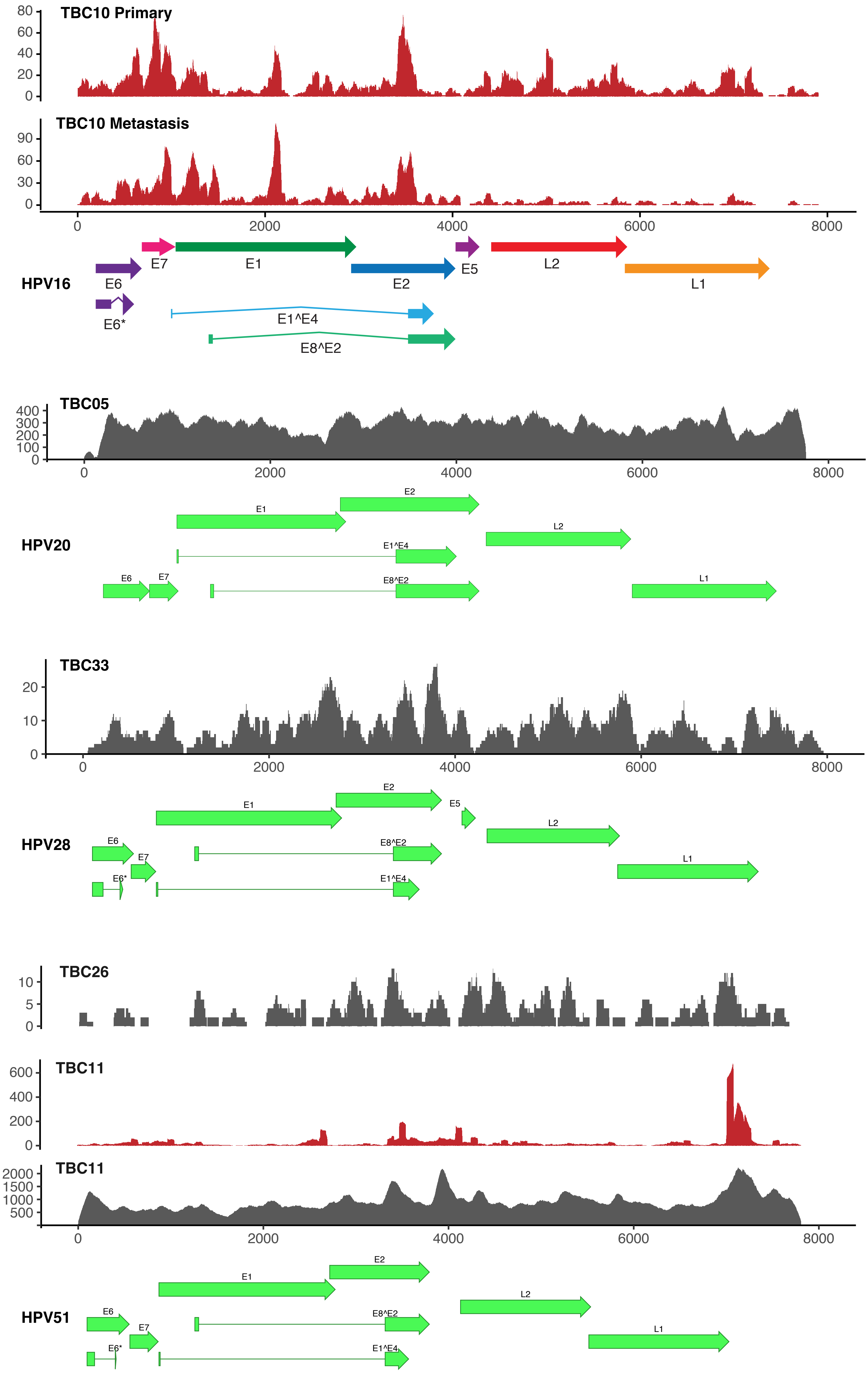

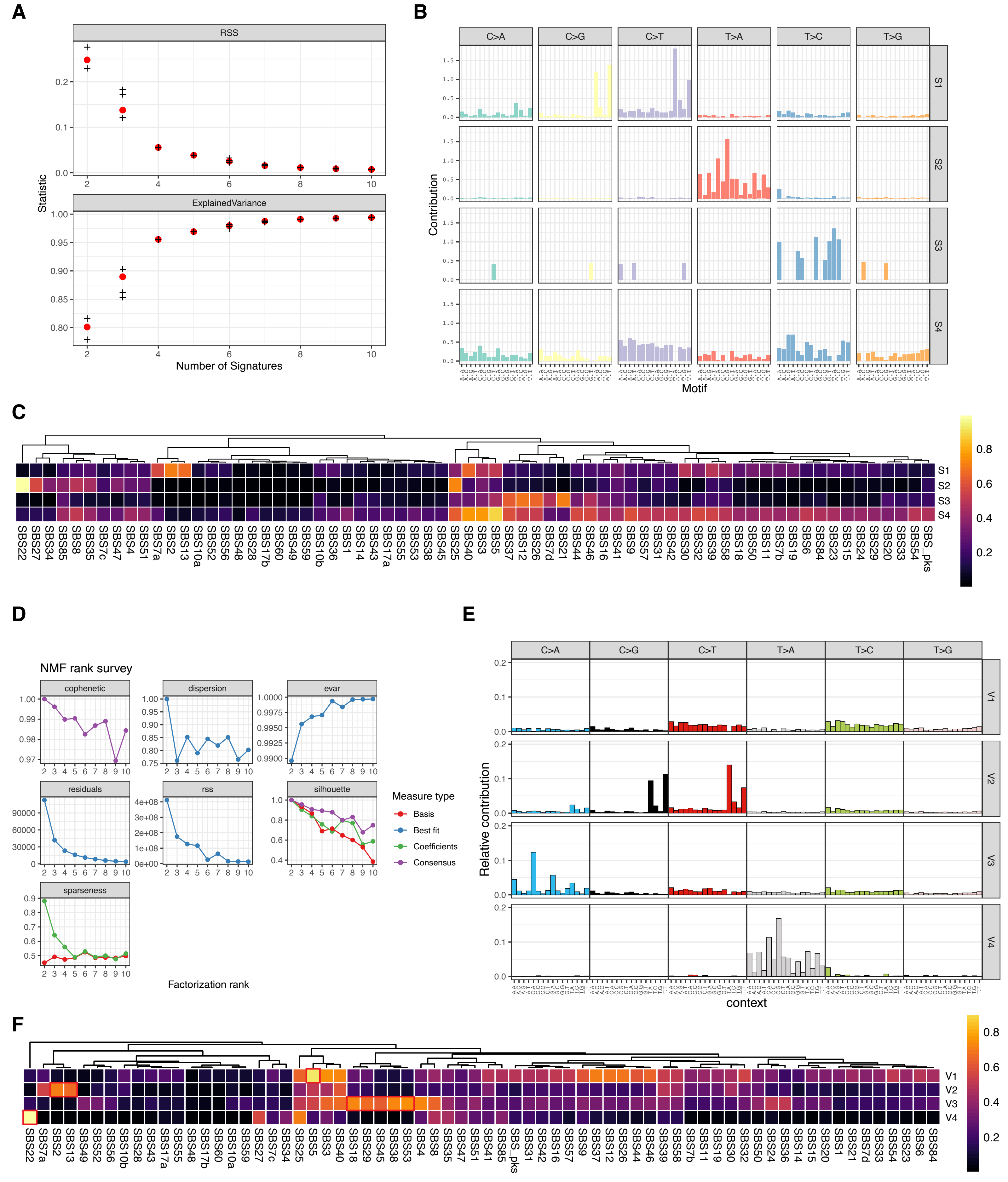

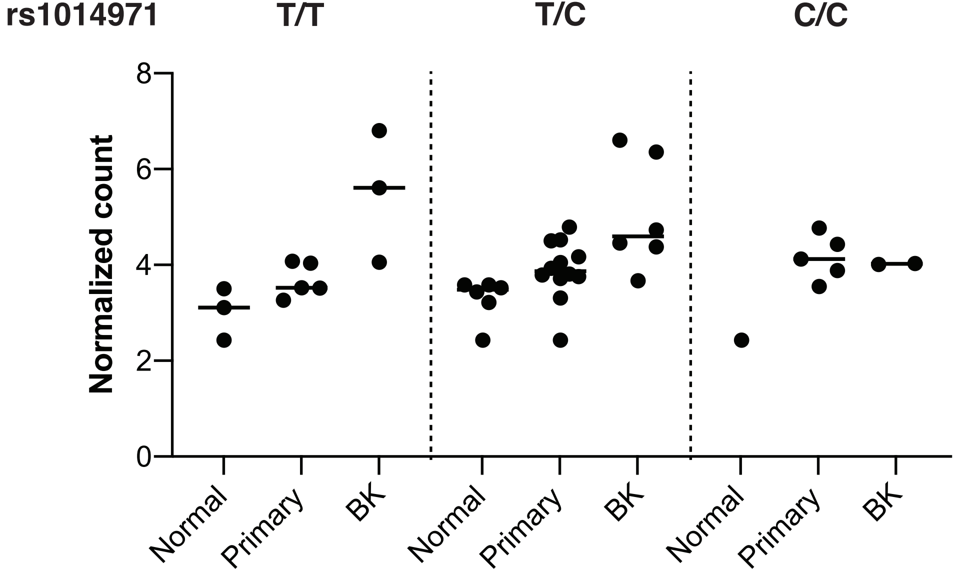
